# An e-Leadership Training Academy for Practicing Clinicians in Primary Care and Public Health Settings

**DOI:** 10.1101/2020.04.17.20068999

**Authors:** Erin Sullivan, Dena Moftah, PaMalick Mbye, Taylor Weilnau, Jonathan N. Tobin

**Affiliations:** Harvard Medical School, Center for Primary Care, Boston, MA; Clinical Directors Network, Inc. (CDN), New York, NY; The Rockefeller University Center for Clinical and Translational Science, New York, NY

## Abstract

**Problem:** There is a lack of leadership training in health care despite it being an essential competency for providers to deliver accessible, high quality healthcare and navigate a continually changing system. The barriers to adding leadership development to the various stages of medical training are numerous. A specific barrier is the lack of access to resources for this training. This group aimed to tackle this barrier within post-graduate medical education and training through their e- Leadership Academy.

**Approach:** The e-Leadership Academy was developed as a partnership between the Harvard Medical School Center for Primary Care and Clinical Directors Network, Inc. (CDN). The result of the collaboration was a virtual leadership academy, offered over a 10-month period that covered the fundamental concepts and skills for leading within a clinical practice. The audience for this program were clinicians and staff of community health centers and health departments in the United States.

**Outcomes:** For the results of this article, primary outcome analysis was of participant responses to both formative and summative evaluations that took place throughout and at the end of the course. Results were used to assess course quality, participant satisfaction, participant engagement, and provide data about future offerings that would be useful to the target audience.

**Next Steps:** The group proposes future training programs could measure the changes in the behavior of teams and clinical outcomes utilizing expanded evaluations. Proposed plans for expansion of the e- Leadership Academy include developing additional modules and the potential integration of an in-person component.

## Problem

Healthcare has lagged behind many other industries in terms of leadership training despite suggestions that it is an essential competency for physicians to provide accessible, high quality healthcare and navigate a continually changing system ^1,2^. Since 2012, peer-reviewed literature shows a marked increase in papers discussing physician leaders. Studies have also demonstrated that training physician leaders improves physician satisfaction ^3^. Along with an increasing demand for physician leaders, there has been a significant expansion in the number of courses to train physicians and other clinician leaders in this area. This is an encouraging development given that medicine tends to select leaders based on demonstrated clinical excellence or scientific innovation, but these forms of success do not necessarily translate to strong leadership proficiency for clinicians ^4^.

Leadership training in medical schools tends to fall into three categories: 1) dual degree programs (MD/MBA); 2) longitudinal tracks with a leadership focus; and 3) classes that range from a one-off session, and autobiographical soliloquies, to multiple, months-long components. At the medical residency level of training, some residency programs have offered leadership curricula or “tracks.” Post-residency, there are a few specialized leadership fellowships for physicians, and a growing number of continuing education leadership training opportunities for physicians and other clinicians at varied career stages. In most of these examples, trainees opt-in to the leadership development track or course, which increases the likelihood that participants are actively seeking a management or leadership-oriented career. It does not account for those who might become “accidental” or “volunTOLD” leaders at some point in their lifetime and find themselves looking for leadership training to support them in a new, and sometimes unexpected role. ^5^

The barriers to adding leadership development to the various stages of medical training are numerous and well-documented elsewhere ^6,7^. One of these barriers is access to resources for training, where resources are defined as money and time. Clinical Directors Network was founded as a practice-based research network and clinical leadership organization, based on the principal of peer-to-peer learning and support, originally presenting clinical leadership training through conferences and workshops in the 1990s ^5^, and then migrating to online CME-accredited webcasts since 2000. We aimed to tackle this barrier to leadership development within post-graduate medical education and training through our e-Leadership Academy.

## Approach

The e-Leadership Academy was developed as a partnership between the Harvard Medical School Center for Primary Care and Clinical Directors Network, Inc. (CDN) in 2018, after a successful 3- webinar pilot series in 2017-2018.

Clinical Directors Network, Inc. (CDN) is a not-for-profit clinician membership organization, practice-based research network (PBRN) and clinician training organization. CDN provides peer-based activities for clinicians practicing in low income, minority, and other underserved communities. CDN’s goal is to translate clinical research into clinical practice for the enhancement of health equity and improvement of public health. CDN offers over 900 online CME-accredited programs in collaboration with other institutions designed to engage and encourage clinical leaders and team members to critically examine their delivery of care.

The Harvard Medical School Center for Primary Care works to strengthen health care through the transformation of systems, teams, and leaders. Developing leaders within primary care has been a core part of the Center’s mission since its inception in 2011. The Center’s leadership development portfolio includes programs that train leaders at various career stages, from medical students through health system executives. The partnership with CDN presented an opportunity for the Center to re-package and digitize its basic leadership development content for an online and national audience of health center and health department clinicians and staff.

The result of the collaboration was a 100% virtual leadership academy, offered over a 10-month period that covered the fundamental concepts and skills for leading within a clinical practice. CDN’s audience for this program was mainly Federally Qualified Health Centers (FQHCs), other community health centers and health departments across the U.S. It is unlikely that these typically resource-constrained organizations would be able to afford sending an entire team to an in-person leadership development program, but the lower cost of distance learning makes it more feasible to support teams in attending a monthly webinar.

Given the interprofessional nature of health care teams, we aimed to be as inclusive as possible in participant recruitment. The course was promoted as appropriate for current or aspiring clinical leaders from a variety of backgrounds: physicians, nurse practitioners, physician’s assistants, nurses, pharmacists, administrators, and social workers. Training is often ineffective if only one individual is trained ^8^, so we encouraged entire teams to register and participate together in the course and a multi-registrant discounted pricing structure was designed to facilitate team participation. Many participants used their CME budgets to support team participation.

Studies on clinician leadership development lack a consensus on the core skills and competencies clinicians need to lead, such as problem-solving skills and communication skills, high degrees of emotional intelligence and a deep understanding of topics like quality improvement, payment, and health systems ^9^. We opted to design the curriculum for the e-Leadership Academy based on the Center for Primary Care’s in-person Leadership Academies that are held in the greater Boston area. The overarching goal was for participants to learn the fundamental concepts and skills for leading oneself, a team, and change within clinical practice.

We created a 10-month series comprised of a monthly 90-minute webinar. The 10 sessions were divided into three specific modules, each with their own key objectives:

1. Leadership Concepts (sessions 1-3): Participants will understand fundamental concepts related to leadership and identify their own specific leadership style.
2. Leading Teams (sessions 4-7): Participants will learn how to build and maintain an effective team; develop strategies for hiring and performance evaluation; and optimize communication and have difficult conversations.
3. Leading Change (sessions 8-10): Participants will evaluate frameworks for leading change; assess challenges in responding to change due to the external environment or internal initiatives; and build strategies for resilience and joy in work in a constantly changing healthcare landscape.

(For full curriculum, see Table 1)

**Table 1.** Course Curriculum Outline.

| # | Title | Description |
| --- | --- | --- |
| **Leadership Concepts** |  |  |
| 1 | **What do leaders do?** | During this session, participants will review the difference between leaders and managers, and understand a leader's key activities and qualities. Participants will dive deeply into one of the core competencies of leadership, setting a vision, and understand how vision affects organizations. |
| 2 | **Understanding different leadership styles** | This session will review the six styles of leadership, the effects of each style, and when to use each style. Participants will also have an opportunity to reflect on his/her own leadership style as well as other styles that they have observed in the workplace. |
| 3 | **"The Es" of Leadership** | This session will focus on describing the three Es of leadership: emotional intelligence, emotional agility and emotional courage. Participants will review strategies to improve these Es as a leader, understand how the Es connect to each other, and discuss why the Es help make stronger leaders. |
| **Leading Teams** |  |  |
| 4 | **Building a team** | This session will explore key elements of teams in healthcare settings and will set the stage for the three sessions that follow in this module. In this session, participants will review what a team is and what makes a team effective. |
| 5 | **Hiring strategies** | This session will start with the first step in building a team: the hiring and onboarding process. Participants will identify successful strategies for developing job descriptions, interviewing candidates, negotiating an offer and incorporating a new hire into your team. |
| 6 | **Optimizing the team** | This session aims to give you some concrete strategies for day-to-day management of your team, including managing meetings, giving feedback and team problem-solving. |
| 7 | **Managing conflict and difficult conversations** | This session will focus on strategies for managing and resolving conflict. It will also examine the elements of a difficult conversation and discuss a framework for holding those conversations. |
| **Leading Change** |  |  |
| 8 | **Quality Improvement: Tools and techniques** | This session will focus on a review of the quality improvement (QI) tools and techniques for process improvement. Participants will discuss how QI supports change within their teams and organizations. |
| 9 | **Power, Persuasion, and Influence** | This session will focus on leading change using power, persuasion and influence. Participants will discuss how to |
|  |  | use persuasion and influence when they do not have a high degree of power or authority. |
| 10 | Leading a Joyful Practice | This final session will discuss how to lead in a way that supports joy, purpose and meaning in work in an ever-changing and challenging healthcare environment. |

Faculty were affiliated with the Center for Primary Care. Each session was structured with 50-60 minutes of content delivery by faculty presenters, which included audience engagement activities via polls and chat boxes; chat box responses were summarized using frequency-based word clouds. A thirty-minute question and answer session followed content delivery. Participants entered questions into a chat box during the content presentation, and faculty answered questions presented by the CDN moderator. Webinars were made available on-demand to all participants so those unable to attend the live sessions were able to attend at their convenience. The complete online course was accredited for up to 15 CME/CNE credits (1.5 credits awarded per session), and carries enduring CME/CNE accreditation so new participants can register and view the full course on-demand and asynchronously.^1^

## Outcomes

Assessment of the e-Leadership Academy included both formative and summative evaluations. In order to receive CME credit for each session, participants were required to complete a 5-question quiz after attending, or viewing on-demand, each monthly session. Additionally, for each session, we included 3-4 questions related to participant satisfaction, feedback on course content, and feedback on the faculty presenter. For the results of this article, we primarily analyzed the participant responses of the final course survey. The survey was conducted at the conclusion of the course to assess course quality, participant satisfaction, and provide data about future offerings that would be useful to our target audience.

### Participation and Engagement

Two hundred seventy six people from 36 states/territories participated, the top three states being California (16.7%), New York (16.3%), and Maryland (9.1%). The majority (72%) registered as part of teams, with team sizes of 2-9 participants (45%), 10-20 participants (19%), or 21+ participants (8%). A total of 15 different occupations were represented, with physicians (46%) and administrators (18%) being the largest categories. The trainees were drawn from 198 organizations (including medical, science, public health and advocacy organizations). Participant engagement was measured by use of attention scores. Attention scores are captured passively and defined as the percent of time the webinar window was the active screen on the participant’s internet-connected device. Participants displayed a cumulative average of 88% attention to the course sessions, with a total of 85,364 active minutes of attention to the course programming (see Table 2).

**Table 2.** Participant Attendance and Engagement.

| Session Name | Cumulative Minutes Viewed | *Average Attention Score |
| --- | --- | --- |
| Session 1: What Do Leaders Do? | 8,616 | 93% |
| Session 2: Understanding Different Leadership Styles | 11,026 | 98% |
| Session 3: “The Es” of Leadership | 9,598 | 88% |
| Session 4: Building a Team | 8,385 | 92% |
| Session 5: Hiring Strategies | 9,903 | 99% |
| Session 6: Optimizing the Team | 7,980 | 86% |
| Session 7: Managing Conflict and Difficult Conversations | 8,224 | 90% |
| Session 8: Quality Improvement: Tools and Techniques | 4,914 | 65% |
| Session 9: Leading a Changing and Diverse Workforce | 9,278 | 90% |
| Session 10: Leading a Joyful Practice | 7,436 | 83% |
| **TOTAL** | **85,364** | **88%** |

\*Attention scores are calculated by Zoom and defined as the percent of time the Zoom webinar window was the active screen on the participant’s device

### Satisfaction data

Qualitative data from participant evaluations were collected on a five-point Likert scale ranging from 1=Poor to 5=Excellent and analyzed both during, and at the conclusion, of the course. In the final evaluation, 96% of participants reported that they were either “Very Satisfied” or “Satisfied” with the course; 92% said that they were likely to recommend it to a colleague/friend, and 88% indicated that the course content will help their daily practice. The following aspects of the course were rated as the most valuable: leadership concepts and theories, content related to the work environment (team building, burnout, culture, workforce), and managing difficult situations. Respondents rated the subject matter organization and delivery on a five-point Likert scale ranging from 1=Poor to 5=Excellent and 99% rated the subject matter organization and delivery of the course as “Excellent” or “Good”; and 96% of respondents rated the quality of the course as “Excellent” or “Good.”

In the final survey, we asked participants about modalities that best support their professional development goals. The results confirm that for our target audience, over 50% prefer an entirely virtual course (see Figure 1). We also learned that in a virtual format, our audience felt connected to each other via the “chat” feature and the Q&A segment of each session, where they reported being able to share insights or learn from other organizations. Participants also found the course accessible, commenting that “once a month is easier to keep up with for someone with a busy schedule” and “having access to the recordings afterwards was a great feature for the meetings.”

**Figure 1.**
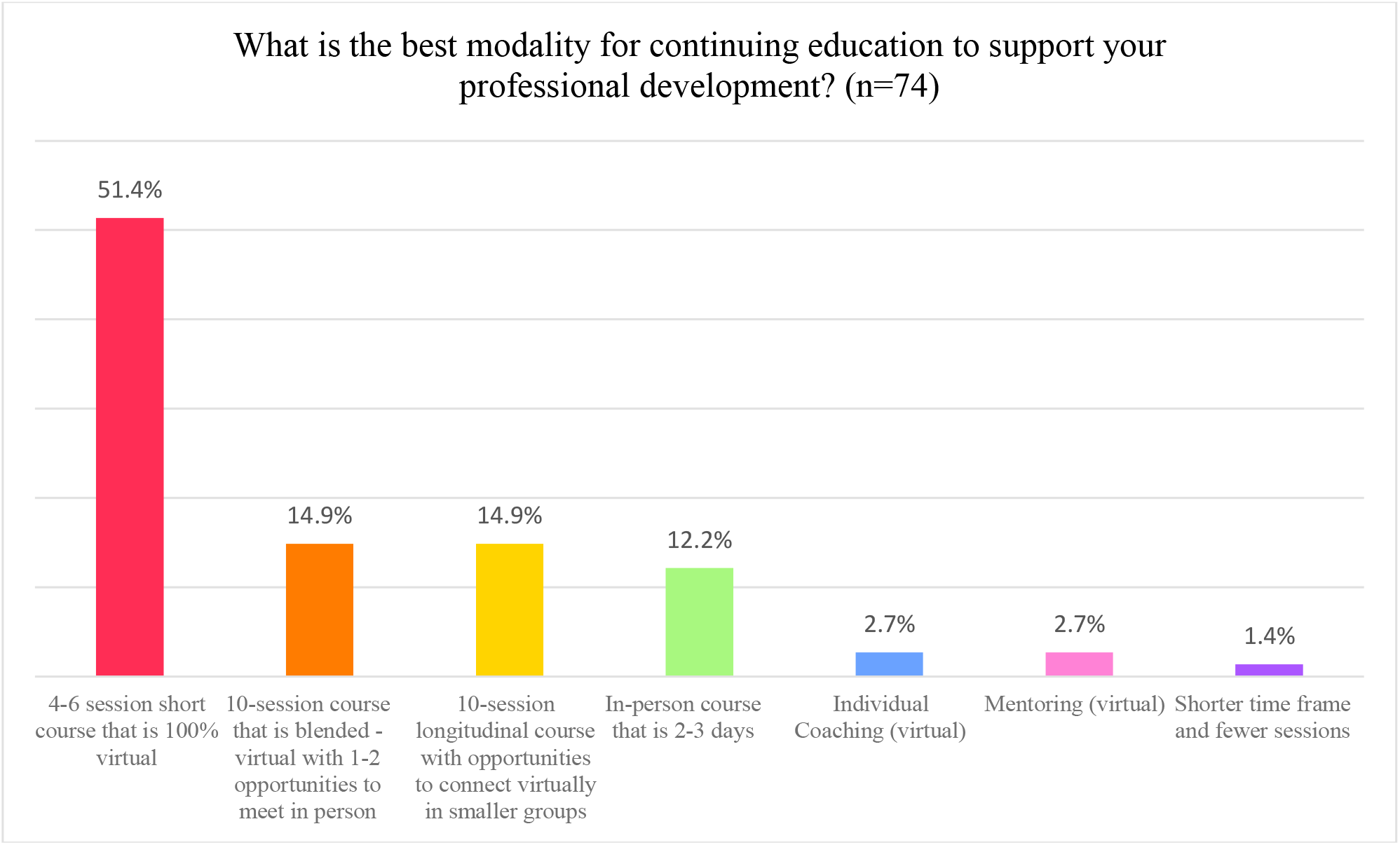
Preferred Modality for Continuing Education.

## Next Steps

The Harvard/CDN e-Leadership Academy focuses on delivering relevant content so that our clinical audience can increase its knowledge. We acknowledge that acquiring new knowledge and skills does not necessarily translate into measurable action. We did not collect any independent measures of implementation or impact of the new content in clinical practice. Future training programs could measure the changes in the behavior of teams (process) and the clinical impact (outcomes) perhaps using EHR data. Nevertheless, the course was designed to provide learners with an opportunity to assimilate what they are learning, try new ways of leading and managing, and reflect on those actions or experiments. For that reason, learners in the proposed Year 2 of the e-Leadership Academy will view recorded webinars from Year 1, and complete an activity that allows them to directly apply the learning principles into their practice settings during the webinar. Then, the cohort will attend bi-monthly office hours, which are structured to increase participant interaction and engagement with faculty and other learners, and create a space to reflect and seek feedback from faculty and peers. This increases the potential audience and maximizes demands of additional time of the faculty by focusing those interactions on application exercises of the principles in the “flipped classroom” ^10^

Future plans for expansion of the e-Leadership Academy include developing additional modules, or short courses, which build on the initial curriculum and provide more advanced, in-depth content. It would be ideal to identify a way to position the current program as a blended, longitudinal program with an in-person component, such as at the beginning or end of the series. An in-person component would allow the curriculum to include more interactive activities, such as role plays and simulations, in which participants can practice leading and receive real-time feedback from peers. An in-person option would also strengthen the connections between participants and leverage CDN’s long-standing peer support network that remains accessible even beyond the conclusion of the program.

## Data Availability

The data that support the findings of this study are available from the co-first authors, upon reasonable request.

## Footnotes

1 www.CDNetwork.org/HarvardLeadership

